# The relationship between gait asymmetry and stability in people with sub-acute stroke

**DOI:** 10.64898/2026.03.16.26348505

**Authors:** Rowan Staines, Kara Patterson, David Jagroop, Elizabeth L Inness, Avril Mansfield

**Affiliations:** KITE-Toronto Rehabilitation Institute, University Health Network, 550 University Avenue, Toronto, ON, Canada, M5G 2A2; Department of Kinesiology and Health Sciences, University of Waterloo, Waterloo, ON, Canada, N2L 3G1; Department of Physical Therapy, University of Toronto, 160-500 University Avenue, Toronto, ON, Canada, M5G 1V7; Evaluative Clinical Sciences, Hurvtiz Brain Sciences Program, Sunnybrook Research Institute, Toronto, ON, Canada, M4N 3M5

**Keywords:** Stroke, Gait, Postural balance, Biomechanical phenomena, Motion capture, Accidental falls

## Abstract

**Background:** People with stroke often walk with temporal asymmetry; which is related to increased fall risk. The purpose of this study was to determine the relationship between temporal gait asymmetry and mechanical stability among people with sub-acute stroke.

**Methods:** Thirty-one people with sub-acute stroke (<6 months post-stroke) completed six walking trials in a biomechanics laboratory. Three-dimensional motion capture was recorded. Swing symmetry was calculated as a ratio of swing time on the more affected limb divided by swing time on the less affected limb. Mechanical stability was the minimum margin of stability, relative to the medial and lateral borders of the stance limb, during the single support phase of the gait cycle. Multiple linear regression was used to determine the relationship between swing symmetry and mechanical stability, controlling for step width and walking speed.

**Results:** There was a significant negative relationship between swing symmetry and lateral margin of stability on the less affected side (p<0.0001) and medial margin of stability on the more affected side (p=0.023). That is, as swing symmetry increased, the extrapolated centre of mass tended to be closer to the lateral border of the less affected side and farther from the medial border of the more affected side.

**Conclusion:** Gait asymmetry could, in part, result from a strategy to compensate for poor balance control on the more affected side. Alternatively, reduced lateral margin of stability on the less affected side among asymmetric participants indicates instability in this direction and could increase the risk for falling.

## INTRODUCTION

People with stroke are at an increased risk of falling in all stages of stroke recovery [1,2], and many falls happen during walking [2]. While regaining walking function is among patients’ top goals of rehabilitation post-stroke [3], and gains are made with rehabilitation, most people with chronic stroke have persistent gait deviations including temporal gait asymmetry [4,5]. Temporal gait asymmetry (TGA) is an interlimb difference in temporal gait variables, such as stance or swing time, and is typically observed as a longer swing phase of the affected limb compared to the unaffected limb [6–8]. Asymmetry provides some insight into the control of walking [4]. TGA is likely a result of a combination of pure neurological impairment following stroke and compensation strategies to reduce time spent on the affected leg during gait [9]. Increased TGA is associated with impaired balance [10], reduced weight bearing capacity on the paretic limb [11], and increased fall risk in people with stroke [12].

Mechanical stability is characterized by the relationship between the centre of mass (COM) and the base of support (BOS) [13]. Maintaining stability during bipedal walking is a complex task, as the COM is generally outside of the BOS during most of the single-support phase of walking. During walking, the COM oscillates between the feet [14,15] and mediolateral stability is actively maintained by the nervous system [16,17]. Asymmetric walking means that people with stroke often maintain their centre of mass toward their unaffected side [18]. The margin of stability (MOS) quantifies how close an inverted pendulum is to falling [19] and can be used to measure mechanical stability during walking [17]. Previous research has shown that people with stroke often have a larger lateral margin of stability on their affected side compared to their unaffected side [20], indicating greater stability towards the affected side compared to the unaffected side.

Although TGA and mediolateral MOS are both related to measures of dynamic balance [10,21,22], few studies have examined the relationship between gait asymmetry and stability during walking. One study by Buurke et al., (2019) [23] found that decreased single support time is related to increased mediolateral (ML) margin of stability in healthy young adults. It is possible that the decreased single support time on the affected leg, seen in post-stroke asymmetric gait, is a strategy to maximize lateral stability on the affected leg. The purpose of the current study is to describe the relationship between gait asymmetry and mechanical stability in people with sub-acute stroke. We hypothesized that swing time asymmetry will be negatively correlated with medial margin of stability on the more affected limb and lateral margin of stability on the less affected limb.

## METHODS

### Design and participants

This is a cross-sectional analysis of data from an assessor-blinded pilot randomized control trial of reactive balance training in sub-acute stroke (NCT04219696) [24] that took place at the Toronto Rehabilitation Institute. Participants were community dwelling adults with subacute stroke (<6 months post-stroke). All participants were able to stand independently for >30 s and walk without a gait aid for at least 10 m. Participants were excluded if they had any contraindications to reactive balance training [24,25]. Written informed consent was obtained from all participants prior to data collection. The study was approved by the Research Ethics Board of the University Health Network (Study ID: 19–6001).

### Cohort descriptors

Demographic, stroke information and medical history were extracted from participants’ hospital charts or directly from the participant. Participant height and body mass were measured. The following assessments were conducted by a trained research assistant: National Institutes of Health Stroke Scale [26]; Chedoke-McMaster Stroke Assessment [27] foot and leg scores; mini-Balance Evaluation Systems Test [28]; and the Activities-specific Balance Confidence scale [29]. For participants with bilateral strokes, the more affected side of the body was the side with lower Chedoke-McMaster Stroke Assessment Scores.

### Overground gait assessment

Data were collected in a biomechanics laboratory at the Toronto Rehabilitation Institute. The laboratory includes a 6 m x 3 m movable platform equipped with 9-12 motion capture cameras (Vicon Mx 40+ and MX F20 or Vicon Vero 2.2, Vicon Motion Capture Systems Ltd., Oxford, UK). Participants were outfitted with 45 single reflective markers on the following anatomical landmarks bilaterally: tip of big toe, 2^nd^ metatarsal-phalangeal joint, 3^rd^ metatarsal phalangeal joint, calcaneous, lateral malleolus, medial malleolus, lateral femoral condyle, medial femoral condyle, greater trochanter, acromion process, lateral humeral epicondyle, medial humeral epicondyle, radial styloid process, and ulnar styloid process. Marker clusters were also placed on the head, upper arms, thighs, shanks, chest, and pelvis. Participants wore a safety harness attached to a robotic gantry overhead to prevent falls, enabling them to move freely within the space without providing body weight support.

Before the test trials, participants performed 3 familiarization walking trials to allow them to become accustomed to walking within the space while equipped with the instrumentation and wearing the safety harness. Participants then performed 10 walking trials on the movable platform at a self-selected pace. The platform moved forward or backward suddenly during four of the trials; the other 6 trials were unperturbed. The perturbed and unperturbed trials were presented in a pseudorandom order, to ensure the participants could not anticipate the perturbations. The six unperturbed trials were analyzed in the current study. The two steps (one on each foot) taken closest to the middle of the platform during each unperturbed trial were used in the analysis.

### Data processing

Motion capture data were filtered in Visual 3D using a lowpass Butterworth filter with a cut-off frequency of 6 Hz. Heel strike and toe-off events were identified using velocity thresholds of 0.1 m/s on descent for heel strike and 0.1 m/s on ascent for toe off. Swing time was defined as the time between toe-off and heel strike events of the swing leg. Swing time asymmetry was calculated using a symmetry ratio, defined as the swing time of the more affected limb divided by the swing time of the less affected limb. Step width was calculated as the mediolateral distance between the heel markers at heel strike.

Whole body COM was calculated using an 11-segment model (i.e., head and trunk, upper arm, forearm and hand, thigh, shank, foot) based on kinematic and anthropometric data [30]. A head, arms and trunk model was used for 11 trials over three participants due to limited visibility of arm markers. COM velocity in the anteroposterior direction was used to measure gait speed.

The extrapolated centre of mass (*XCOM*) was calculated using Equation 1 [19]:

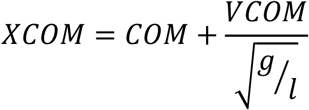

Where *COM* was the position of the COM, *VCOM* was the velocity of the COM, *g* was the gravity constant (9.81m/s^2^), and *l* was the participant’s height multiplied by the greater trochanter height of 0.53 [30] and multiplied by 1.34 [19].

The margin of stability was then calculated using Equation 2 [19]:

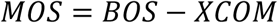

Where the BOS was the boundary of the base of support. MOS was calculated relative to the medial and lateral borders of the stance foot. The medial and lateral borders of the BOS were defined as the position of the toe and 5^th^ metatarsal markers, respectively. A positive MOS represents a situation where the XCOM is within the BOS boundary, whereas the XCOM outside the boundary of the BOS is represented as a negative MOS. Therefore, the margins of stability of the right foot were multiplied by -1 to align with the laboratory coordinate system; this ensured that the MOS was positive when ‘inside’ (i.e., medial to) the lateral border of both the left and right feet. Medial and lateral MOS were calculated during single support phase. We used both the medial and lateral borders of the BOS to calculate medial and lateral MOS separately to better understand the location of the XCOM in relation to the entire BOS during walking.

### Data analysis

Statistical analyses were performed using Stata (Version 18.5, StataCorp LLC., College Station, Texas, USA). Participants with missing data were excluded from the analysis of that variable. For descriptive purposes, participants were split into symmetric and asymmetric groups based on their symmetry ratio; symmetric (0.94-1.06), asymmetric towards less affected side (>1.06), and asymmetric towards the more affected side (<0.94) [7], and cohort descriptors were presented separately for each group. The minimum medial and lateral margins of stability for each single support period were used for analysis. Multiple linear regressions with generalized estimating equations and repeated measures (to account for multiple observations per participant) were used to assess the relationships between minimum lateral MOS of the less affected leg and swing symmetry, and minimum medial MOS of the more affected leg and swing symmetry. Dependent variables in the regression were lateral MOS on the less affected side or medial MOS on the more affected side, and independent variables were swing symmetry, step width, and gait speed (confounding variables). Alpha was 0.05 for all analyses.

## RESULTS

There were 31 participants included in the analysis. Participant characteristics are displayed in Table 1.

**Table 1:**
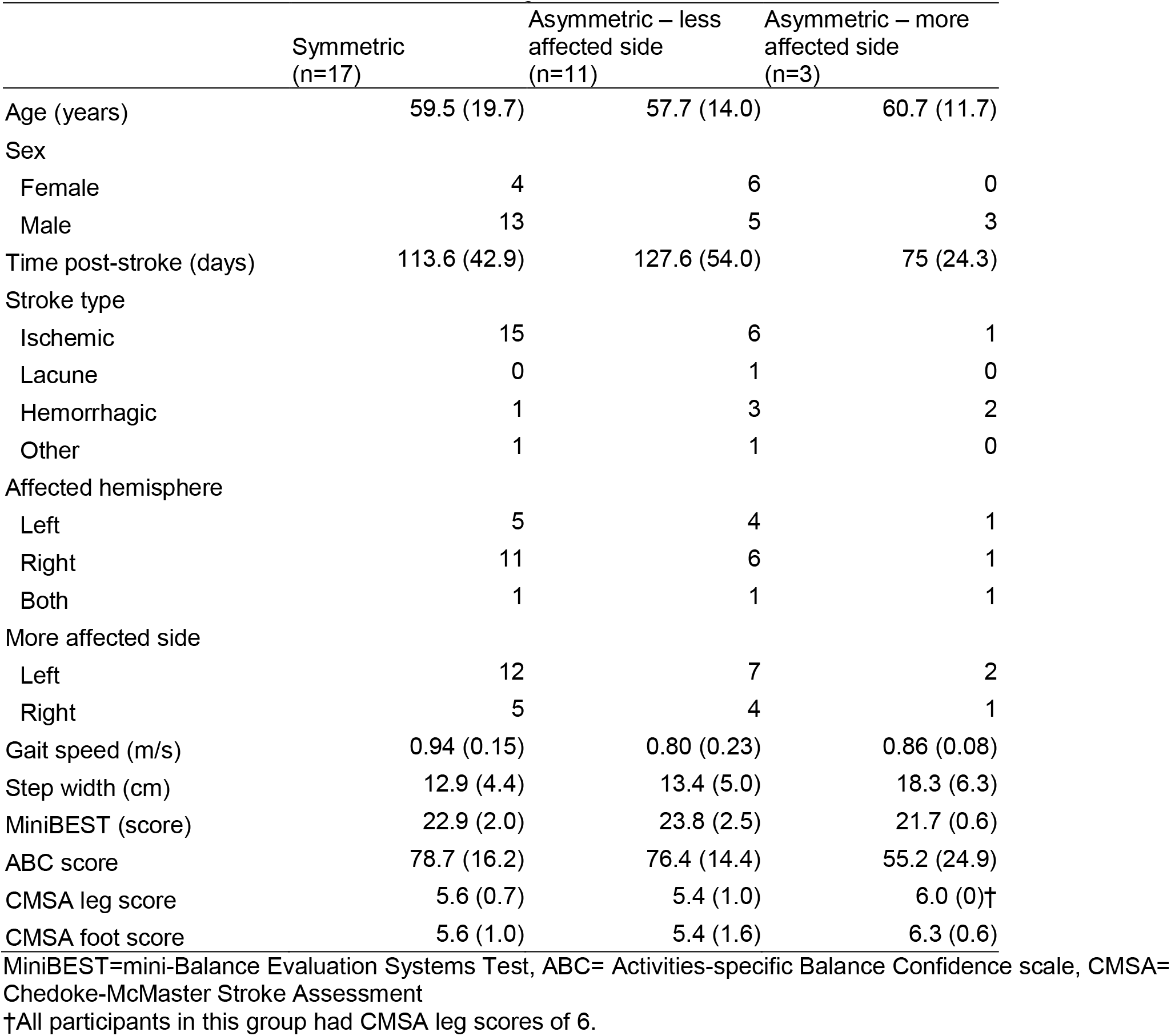
Participant characteristics. Values presented are means with standard deviations in parentheses for continuous or ordinal variables, or counts for categorical variables.

Figure 1 shows an illustrative example of XCOM relative to the BOS for a symmetric and asymmetric trial. Results of multiple linear regressions with generalized estimating equations and repeated measures are shown in Table 2. In support of our hypothesis, there was a significant negative relationship between swing time symmetry and lateral MOS of the unaffected leg (p < 0.0001), and between swing time symmetry and medial MOS of the affected leg (p=0.023) when controlling for step width and walking speed (Figure 2).

**Table 2:** Results of multiple linear regression analyses. Estimates are the slopes for each term in the model, with 95% confidence intervals in brackets.

|  | Estimate | p-value |
| --- | --- | --- |
| MOS-M more affected (cm) |  |  |
| Swing symmetry (ratio) | -4.28 [-7.96, -0.60] | 0.023 |
| Step width (cm) | -0.68 [-0.79, -0.56] | <0.0001 |
| Walking speed (m/s) | -5.31 [-10.22, -0.39] | 0.034 |
| MOS-L less affected (cm) |  |  |
| Swing symmetry (ratio) | -4.21 [-6.04, -2.39] | <0.0001 |
| Step width (cm) | 0.13 [0.07, 0.18] | <0.0001 |
| Walking speed (m/s) | 1.55 [-0.97, 4.07] | 0.23 |

MOS-M= medial margin of stability, MOS-L= lateral margin of stability

**Figure 1:**
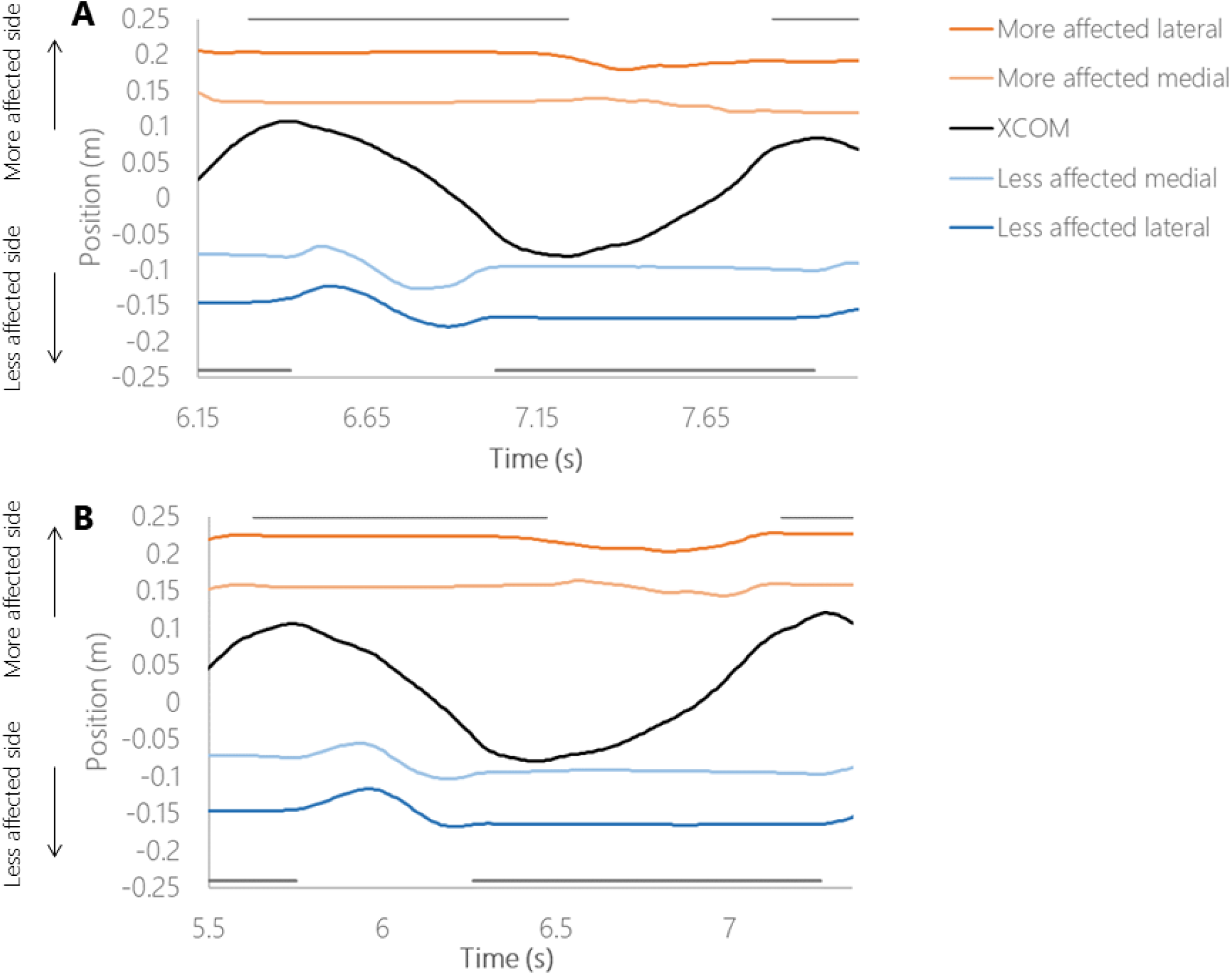
Sample time series. Extrapolated centre of mass (XCOM) and lateral and medial edges of the base of support are plotted for an illustrative trial where the participant was symmetric (Panel A) and asymmetric (Panel B). Stance times for the more affected and less affected limbs are indicated by the grey horizontal lines at the top (more affected) and bottom (less affected) of each figure. The XCOM is farther from the base of support during the asymmetric trial compared to the symmetric trial.

**Figure 2:**
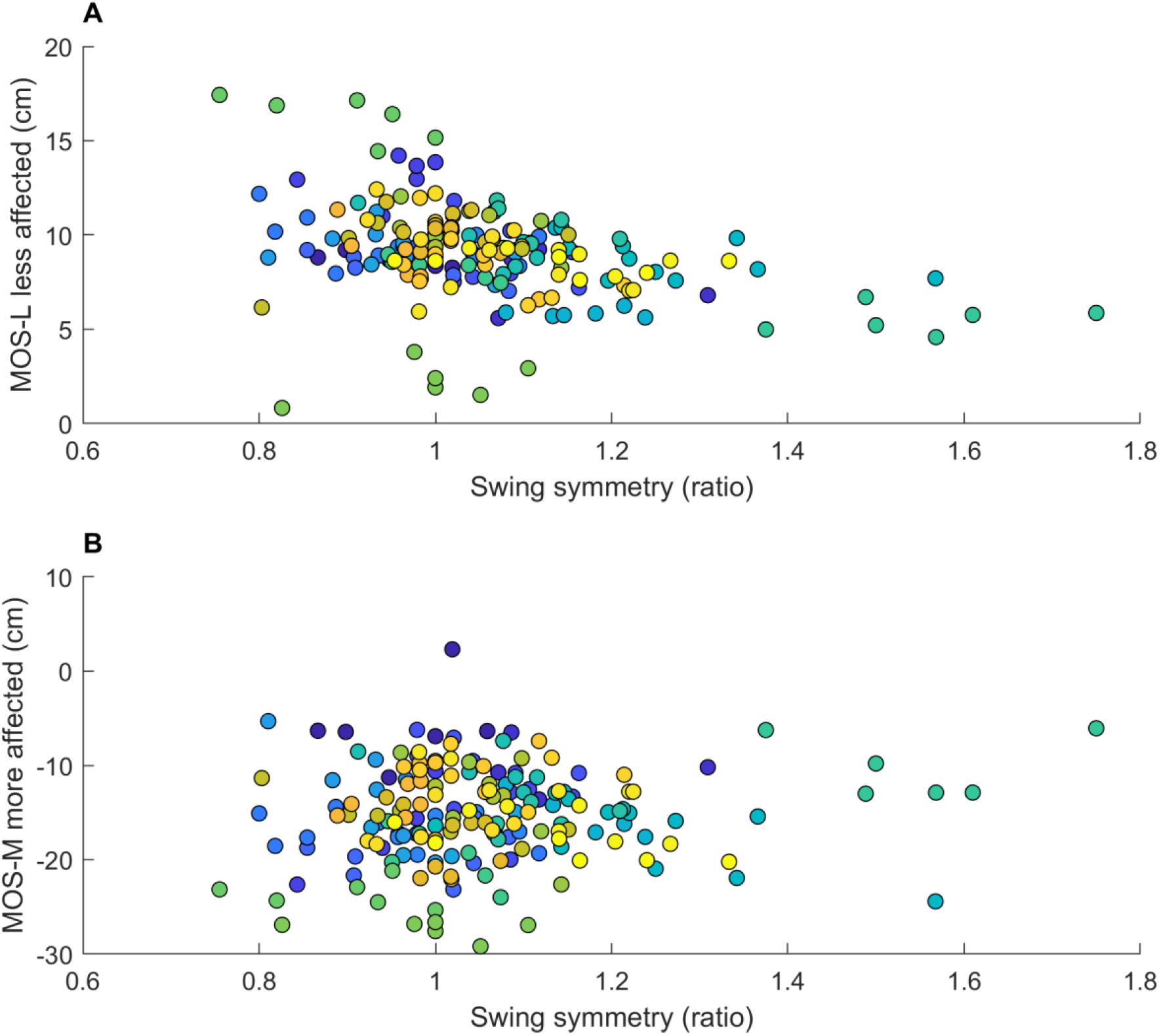
Relationship between swing symmetry and margin of stability. Margin of stability toward the lateral edge of the base of support (MOS-L; panel A) and medial edge of the base of support (MOS-M; panel B) are plotted against swing symmetry. Each point represents a single trial for each participant; points of the same colour are from the same participant.

## DISCUSSION

The aim of this cross sectional analysis was to describe the relationship between gait asymmetry and stability during walking in people with sub-acute stroke. We found that swing time symmetry was negatively related to lateral margin of stability on the less affected leg and to medial MOS on the more affected leg. Specifically, when swing time asymmetry is increased, with greater time spent on the less affected leg, the COM remains closer to the lateral border of the less affected foot and farther from the medial border of the more affected foot. The relationship between swing symmetry and MOS was independent of step width and gait speed. Previously, authors have speculated that gait asymmetry could, in part, result from a strategy to compensate for poor balance control [4], or that instability could be negative consequence of gait asymmetry [5]. This study provides support for these speculations.

The results from the current study support previous research that people with stroke often walk with a larger lateral MOS on the more affected leg compared to the less affected leg [20]. Our study revealed that this characteristic is more prominent in people with greater temporal gait asymmetry. The results of the current study indicated that, while reduced time spent on the more affected leg is related to a decreased lateral margin of stability on the less affected leg, it is simultaneously related to an increased lateral margin of stability on the more affected leg. It is possible that the reduced time spent on the more affected leg may be a strategy to maintain lateral stability on the more affected leg, however in doing so, the lateral margin of stability on the less affected side may be compromised. People with stroke typically prefer to respond to balance perturbation with the less affected leg and may be unable to respond to a loss of balance with the more affected leg [31]. Therefore, a loss of balance towards this less affected side may pose a specific challenge for people with stroke. The apparent strategy to maintain stability by shifting weight towards the less affected side when walking may, paradoxically, actually increase fall risk by hindering their ability to make the preferred reactive step with their less affected side

According to the inverted pendulum model, step width is closely related to lateral MOS, where wider steps are related to a greater MOS [15]. Results from the current study are consistent with this, as there was a positive relationship between step width and lateral MOS of the less affected limb. Conversely, step width was negatively related to medial MOS of the more affected foot. These findings suggest that a wider step may be a response to low mechanical stability when the XCOM moves medially during single support. These findings are in agreement with Sivakumaran et al (2018), who found that medial MOS was lower when preceding wider than usual steps [33].

A few limitations of the current study should be acknowledged. First, the range of asymmetry among participants in the current study did not capture individuals with severe asymmetry, as defined by an asymmetry ratio greater than 1.5 [3]. To participate in the larger study, participants had to be able to walk at least a short distance without a gait aid; therefore, we likely necessarily excluded participants with more severe gait impairments from the study. Consequently, the results may not be generalizable to people with severe gait asymmetry. Second, this study involved participants with sub-acute stroke and there is evidence to suggest that gait asymmetry is worse in the chronic stage of stroke recovery [37]. Future studies should investigate the relationship between gait asymmetry and stability in people with chronic stroke.

## Conclusion

This study describes how gait asymmetry relates to mechanical instability during walking for individuals with subacute stroke. A clearer understanding the relationship between gait asymmetry and stability may be useful to inform stroke rehabilitation approaches. Previous work reports that gait asymmetry has been resistant to intervention [34]. Thus, future studies should explore interventions to improve stability, such as targeting ankle moment control and step placement control [35,36].

## Data Availability

Participants did not consent to sharing data outside the research team.

## Acknowledgements

We thank Abigail Morton and Sarah Thompson for their assistance with data collection.

## REFERENCES

[1] F.A. Batchelor, S.F. Mackintosh, C.M. Said, K.D. Hill, Falls after stroke, Int J Stroke. 7 (2012) 482–490.

[2] A. Mansfield, E.L. Inness, W.E. McIlroy, Stroke, in: B.L. Day, S.R. Lord (Eds.), Handbook of Clinical Neurology: Balance, Gait, and Falls, Elsevier BV, San Diego, 2018: pp. 205–228. https://www.sciencedirect.com/handbook/handbook-of-clinical-neurology/vol/159/suppl/C.

[3] J. Evensen, H.L. Soberg, U. Sveen, K.A. Hestad, J.L. Moore, B.A. Bronken, Individualized goals expressed by patients undergoing stroke rehabilitation: an observational study, J Rehabil Med. 56 (2024) jrm15305. 10.2340/jrm.v56.15305.

[4] K.K. Patterson, I. Parafianowicz, C.J. Danells, V. Closson, M.C. Verrier, W.R. Staines, et al., Gait asymmetry in community-ambulating stroke survivors, Arch Phys Med Rehabil. 89 (2008) 304– 310.

[5] K.K. Patterson, A. Mansfield, L. Biasin, K. Brunton, E.L. Inness, W.E. McIlroy, Longitudinal changes in poststroke spatiotemporal gait asymmetry over inpatient rehabilitation, Neurorehabilitation and Neural Repair. 29 (2015) 153–162. 10.1177/1545968314533614.

[6] K.K. Patterson, W.H. Gage, D. Brooks, S.E. Black, W.E. McIlroy, Evaluation of gait symmetry after stroke: a comparison of current methods and recommendations for standardization, Gait Posture. 31 (2010) 241–245.

[7] E.B. Titianova, I.M. Tarkka, Asymmetry in walking performance and postural sway in patients with chronic unilateral cerebral infarction, J Rehabil Res Dev. 32 (1995) 236–44.

[8] J.C. Wall, G.I. Turnbull, Gait asymmetries in residual hemiplegia, Arch Phys Med Rehabil. 67 (1986) 550–553.

[9] N. Mizuta, N. Hasui, Y. Higa, A. Matsunaga, S. Ohnishi, Y. Sato, et al., Identifying impairments and compensatory strategies for temporal gait asymmetry in post-stroke persons, Sci Rep. 15 (2025) 2704. 10.1038/s41598-025-86167-9.

[10] M.D. Lewek, C.E. Bradley, C.J. Wutzke, S.M. Zinder, The relationship between spatiotemporal gait asymmetry and balance in individuals with chronic stroke, J Appl Biomech. 30 (2014) 31–36.

[11] J. Hendrickson, K.K. Patterson, E.L. Inness, W.E. McIlroy, A. Mansfield, Relationship between asymmetry of quiet standing balance control and walking post-stroke, Gait & Posture. 39 (2014) 177–181. 10.1016/j.gaitpost.2013.06.022.

[12] T.-S. Wei, P.-T. Liu, L.-W. Chang, S.-Y. Liu, Gait asymmetry, ankle spasticity, and depression as independent predictors of falls in ambulatory stroke patients, PLoS ONE. 12 (2017) e0177136. 10.1371/journal.pone.0177136.

[13] S.M. Bruijn, O.G. Meijer, P.J. Beek, J.H. van Dieën, Assessing the stability of human locomotion: a review of current measures, J R Soc Interface. 10 (2013) 20120999. 10.1098/rsif.2012.0999.

[14] A.L. Hof, R.M. van Bockel, T. Schoppen, K. Postema, Control of lateral balance in walking. Experimental findings in normal subjects and above-knee amputees, Gait Posture. 25 (2007) 250– 258.

[15] D.A. Winter, Human balance and posture control during standing and walking, Gait Posture. 3 (1995) 193–214.

[16] T. Fettrow, H. Reimann, D. Grenet, E. Thompson, J. Crenshaw, J. Higginson, et al., Interdependence of balance mechanisms during bipedal locomotion, PLoS ONE. 14 (2019) e0225902. 10.1371/journal.pone.0225902.

[17] A.L. Hof, The “extrapolated center of mass” concept suggests a simple control of balance in walking, Hum Mov Sci. 27 (2008) 112–125. 10.1016/j.humov.2007.08.003.

[18] C.K. Balasubramanian, R.R. Neptune, S.A. Kautz, Foot placement in a body reference frame during walking and its relationship to hemiparetic walking performance, Clinical Biomechanics. 25 (2010) 483–490. 10.1016/j.clinbiomech.2010.02.003.

[19] A.L. Hof, M.G.J. Gazendam, W.E. Sinke, The condition for dynamic stability, J Biomech. 38 (2005) 1–8. 10.1016/j.jbiomech.2004.03.025.

[20] F.B. Van Meulen, D. Weenk, E.H.F. Van Asseldonk, H.M. Schepers, P.H. Veltink, J.H. Buurke, Analysis of balance during functional walking in stroke survivors, PLoS ONE. 11 (2016) e0166789. 10.1371/journal.pone.0166789.

[21] F. Watson, P.C. Fino, M. Thornton, C. Heracleous, R.L. Loureiro, Use of the margin of stability to quantify stability in pathologic gait - a qualitative systematic review, BMC Musculoskelet Disord. 22 (2021) 597. 10.1186/s12891-021-04466-4.

[22] A. Vistamehr, S.A. Kautz, M.G. Bowden, R.R. Neptune, Correlations between measures of dynamic balance in individuals with post-stroke hemiparesis, Journal of Biomechanics. 49 (2016) 396–400. 10.1016/j.jbiomech.2015.12.047.

[23] T.J.W. Buurke, C.J.C. Lamoth, L.H.V. Van Der Woude, A.L. Hof, R. Den Otter, Bilateral temporal control determines mediolateral margins of stability in symmetric and asymmetric human walking, Sci Rep. 9 (2019) 12494. 10.1038/s41598-019-49033-z.

[24] A. Mansfield, E.L. Inness, C.J. Danells, D. Jagroop, T. Bhatt, A.H. Huntley, Determining the optimal dose of reactive balance training after stroke: study protocol for a pilot randomised controlled trial, Bmj Open. 10 (2020) e038073. 10.1136/bmjopen-2020-038073.

[25] A. Mansfield, C.J. Danells, E.L. Inness, K.E. Musselman, N.M. Salbach, A survey of Canadian healthcare professionals’ practices regarding reactive balance training, Physiother Theory Pract. doi:10.1080/09593985.2019.1650856 (2021) 787–800.

[26] L.B. Goldstein, C. Bertels, J.N. Davis, Interrater reliability of the NIH Stroke Scale, Arch Neurol. 46 (1989) 660–662. 10.1001/archneur.1989.00520420080026.

[27] C. Gowland, P. Stratford, M. Ward, J. Moreland, W. Torresin, S. Van Hullenaar, et al., Measuring physical impairment and disability with the Chedoke-McMaster Stroke Assessment, Stroke. 24 (1993) 58–63. 10.1161/01.str.24.1.58.

[28] F. Franchignoni, F. Horak, M. Godi, A. Nardone, A. Giordano, Using psychometric techniques to improve the Balance Evaluation Systems Test: the mini-BESTest, J Rehabil Med. 42 (2010) 323– 331. 10.2340/16501977-0537.

[29] L.E. Powell, A.M. Myers, The Activities-specific Balance Confidence (ABC) Scale, J Gerontol A Biol Sci Med Sci. 50A (1995) M28–34. 10.1093/gerona/50a.1.m28.

[30] D.A. Winter, Biomechanics and Motor Control of Human Movement, 1st ed., Wiley, 2009. 10.1002/9780470549148.

[31] A. Mansfield, E.L. Inness, B. Lakhani, W.E. McIlroy, Determinants of limb preference for initiating compensatory stepping poststroke, Arch Phys Med Rehabil. 93 (2012) 1179–1184. 10.1016/j.apmr.2012.02.006.

